# “Nobody has ever spoken to me about PCD and fertility issues”: Fertility experiences of people with primary ciliary dyskinesia and their family caregivers

**DOI:** 10.1101/2025.02.04.25321417

**Authors:** Leonie D Schreck, Sophie Meyer, Eva SL Pedersen, Yin Ting Lam, Hansruedi Silberschmidt, Sara Bellu, Living with PCD patient advisory group, Sofía C Zambrano, Claudia E Kuehni, Myrofora Goutaki

## Abstract

**Background:** Primary ciliary dyskinesia (PCD) affects fertility in both women and men. To understand the impact and concerns among people with PCD and parents of affected children (family caregivers), we explored how they report their experiences with fertility.

**Methods:** We used qualitative data from a questionnaire on fertility from *Living with PCD*, an international participatory study. In optional open-ended comment fields, participants shared their thoughts and experiences related to fertility. We adopted conventional content analysis and analysed the data inductively.

**Results:** We identified five categories illustrating participants’ experiences with fertility: Challenging experiences of fertility care, PCD-related reproductive concerns, non-PCD-related factors complicating fertility, psychological impact of infertility, and family caregivers as gatekeepers of fertility information.

**Conclusion:** We need enhanced support and standardised reproductive counselling and health care for people with PCD to enable informed decisions on fertility, and to reduce the fertility related concerns and psychological impact faced by many.

**Highlights:**

– People with primary ciliary dyskinesia (PCD) had challenging fertility experiences
– Impact of PCD on pregnancy and disease heritability were causes for concern
– Other reasons not related to PCD but affecting fertility complicated the journey
– Infertility as an emotionally difficult topic strongly impacted participants’ lives
– Family caregivers acted as gatekeepers for fertility information of their children

## Introduction

Understanding the fertility experiences of people with primary ciliary dyskinesia (PCD) and parents of affected children (family caregivers) is important, given the high risk of infertility associated with the condition. PCD is a rare genetic multiorgan disease which impairs the structure and function of motile cilia throughout the body. This ciliary dysfunction mainly leads to chronic upper and lower airway symptoms such as recurrent infections, bronchiectasis, and hearing impairment, significantly impacting the quality of life of those affected (Lucas et al., 2015). Beyond its respiratory manifestations, infertility affects up to 80% of men and 60% of women with PCD (Schreck et al., 2024b). Male infertility is the result of dysmotile sperm and sperm agglutination due to defective cilia in the testes (Sironen et al., 2020). Female infertility, although less well understood, is thought to result from ciliary dysfunction in the fallopian tubes and uterus (Raidt et al., 2015; Newman et al., 2023; Pearson-Farr et al., 2023). Women with PCD also have an increased risk of ectopic pregnancies (Schreck et al., 2024b), likely due to impaired oocyte transport. Recent research highlights a significant need for fertility treatments among both men and women to achieve successful conception (Schreck et al., 2024b). Despite this, referral to fertility specialists is not routinely integrated into PCD care and people with PCD are often dissatisfied with the fertility information provided by their healthcare providers (Schreck et al., 2024a).

To date, there is a significant gap in studies exploring the lived experiences of individuals with PCD and their family caregivers in relation to fertility. Evidence from studies on other conditions associated with infertility suggests that the inability to conceive is often an unexpected and distressing experience, and often a personal tragedy (Cousineau and Domar, 2007; Greil et al., 2010). Among childhood cancer survivors, the uncertainty surrounding their fertility leads to a psychological burden, anxiety and a sense of uncertainty about their own future (Newton et al., 2021). Similarly, adult breast cancer survivors have described the consequences of infertility as “devastating,” resulting in emotional distress, fear, and feeling “broken hearted” (Perz et al., 2014). Alongside these psychological challenges, individuals often face difficulties in communicating with healthcare providers as they navigate their fertility journey (Newton et al., 2021). For women with chronic lung diseases, illness-related challenges often influence reproductive decisions. They often receive conflicting advice regarding the impact of their condition on pregnancy and how pregnancy might affect their health (Williams et al., 2021). Lastly, fertility treatments are known to be physically, emotionally and socially difficult (Cousineau and Domar, 2007; Assaysh-Öberg et al., 2023).

To understand the concerns and impact of fertility problems on people with PCD, we explored how they and their family caregivers report their experiences with fertility. This understanding is essential to identify and address the needs of people living with PCD and is crucial for developing patient-centred resources, support systems, fertility care guidelines, and healthcare services.

## Methods

### Study design and procedure

We analysed free-text data from a questionnaire on fertility and fertility care conducted as part of the *Living with PCD* study (previously named COVID-PCD), an international participatory study involving over 750 participants from 49 countries. *Living with PCD* was established during the coronavirus disease 2019 (COVID-19) pandemic to monitor the impact of COVID-19 on people with PCD. Following the pandemic, the study focused on addressing broader questions raised by individuals with PCD. The topic of fertility was identified as a research priority by both study participants and people with PCD outside the study (Lam et al., 2022). The *Living with PCD* study is led by a research team at the University of Bern, with study decisions made in close collaboration with a study advisory group consisting of members of PCD patient associations worldwide. A detailed study protocol has been published (Pedersen et al., 2021).

*Living with PCD* is an anonymous study open to individuals worldwide who have a confirmed or suspected PCD diagnosis. Participants include adults, adolescents, and family caregivers of children with PCD who complete questionnaires on behalf of their children. Patient associations invited their members to participate by sharing the invitation link via email and on social media. Participants self-registered on the study website, after which they received a questionnaire covering demographic information and details about their PCD diagnosis and symptoms, followed by shorter weekly follow-up questionnaires and occasional questionnaires on special topics. We sent a special questionnaire on fertility and fertility care to all study participants on 12 July 2022 and to all participants who joined later by 8 March 2023. Non-responders received up to three reminders via email. Participants entered data directly into an online database using the Research Electronic Data Capture (REDCap) platform (Harris et al., 2009), hosted by the Swiss medical registries and data linkage centre at the University of Bern, Switzerland. Participants gave informed consent at enrolment and could leave the study at any time by contacting the study team. The ethics committee of the canton of Bern, Switzerland, approved the study (study ID: 2020-00830).

### Data collection/questionnaire

We developed the fertility questionnaire together with fertility specialists, reproductive endocrinologists, and members of the study advisory group. The questionnaire contained mainly closed questions with five open-ended questions, of which the last question invited participants to “Please write any additional comments you may have about fertility in people with PCD or comments to this questionnaire.” The analysis of these additional comments forms the focus of this manuscript. The questionnaire was developed in English, and native-speaking members of the study team translated the questionnaire into French, German, Italian, and Spanish, and two members of the study team or advisory group checked the translations. Responses of participants in languages other than English were translated via DeepL and checked by a native speaking member of the research group. We edited the participants’ quotes for language and clarity (Eldh et al., 2020), and included information on participants’ age, sex, and fertility. To ensure the anonymity of our participants, we report their age in age groups: 0-5 years, 6-12 years, 13-17 years, 18-29 years, 30-39 years, 40-49 years, and ≥ 50 years. We only selected one quote per individual to ensure diverse representations. The results of other parts of the survey are presented elsewhere (Schreck et al., 2024a; Schreck et al., 2024b).

### Participants

Overall, 266 adults, 16 adolescents, and 102 parents or family caregivers of children with PCD completed the fertility questionnaire. Among them, 206 (122 women, 46 men, 4 adolescents, 34 parents) left a comment and were included in this analysis. The adults had a median age of 42 years. Most participants were from the United Kingdom (41; 20%), the United States (33; 16%), or Germany (31; 15%). Two hundred (97%) reported having a PCD diagnosis confirmed by a physician, and six (3%) had a suspected diagnosis. Among 121 adult participants who had already tried to conceive, 38 (31%) had naturally conceived children, 49 (40%) had conceived children with the use of fertility treatments, and 34 (28%) had not been able to conceive a child with or without the use of fertility treatments.

### Analysis

We employed conventional content analysis (Hsieh and Shannon, 2005). This approach is specifically suitable when “existing theory or research literature on a phenomenon is limited” (Hsieh and Shannon, 2005) allowing for researcher immersion with data at hand. We coded data mainly inductively aiming at capturing and reflecting participants’ experiences related to fertility. However, at a later stage, deductive codes relating to care provision and diagnostics helped to organise the findings.

The first author (LDS) undertook the main analyses with support from SM and supervised by MG. LDS is a medical doctor and a PhD student in epidemiology with basic training in qualitative research. Her research is mainly focused on PCD with a particular emphasis on fertility using mainly quantitative methods. SM is a PhD student in public health with a background in social sciences and with in-depth training and experience in undertaking qualitative research. She only had little prior knowledge on the topics of PCD and fertility. MG is a senior researcher and medical doctor with a PhD in epidemiology with extensive experience in qualitative methods and is a PCD expert leading international PCD research collaborations (Goutaki et al., 2021).

LDS read the free text responses several times to gain a general understanding of the information and achieve immersion. By reading every response word by word, she identified initial codes related to participants’ experiences and sorted them into categories, creating an initial coding scheme. SM independently read a selection of open-ended questions, coded them and discussed impressions of participants’ experiences with LDS. In an iterative process, LDS and SM reviewed, compared, and refined the categories by grouping overlapping or similar codes together. If they could not agree on the categories, they went back through the participants’ responses to make sure they had captured what participants expressed and stayed close to the data. They discussed differences in coding and reassessed to reach consensus. The last author, MG, read a random selection of open-ended responses and participated in several meetings to critically reflect upon the analysis procedure and interpretation of findings. We used NVivo 1.7.1 to facilitate data management during analysis.

## Results

We identified five categories illustrating the experiences with fertility of people with PCD and their family caregivers: 1) challenging experiences of fertility care, 2) PCD-related reproductive concerns, 3) non-PCD-related factors complicating fertility, 4) psychological impact of infertility, and 5) family caregivers as gatekeepers of fertility information. Of note, the categories are not to be understood as separate topics as they are interlinked in many instances (Figure 1).

**Figure 1:**
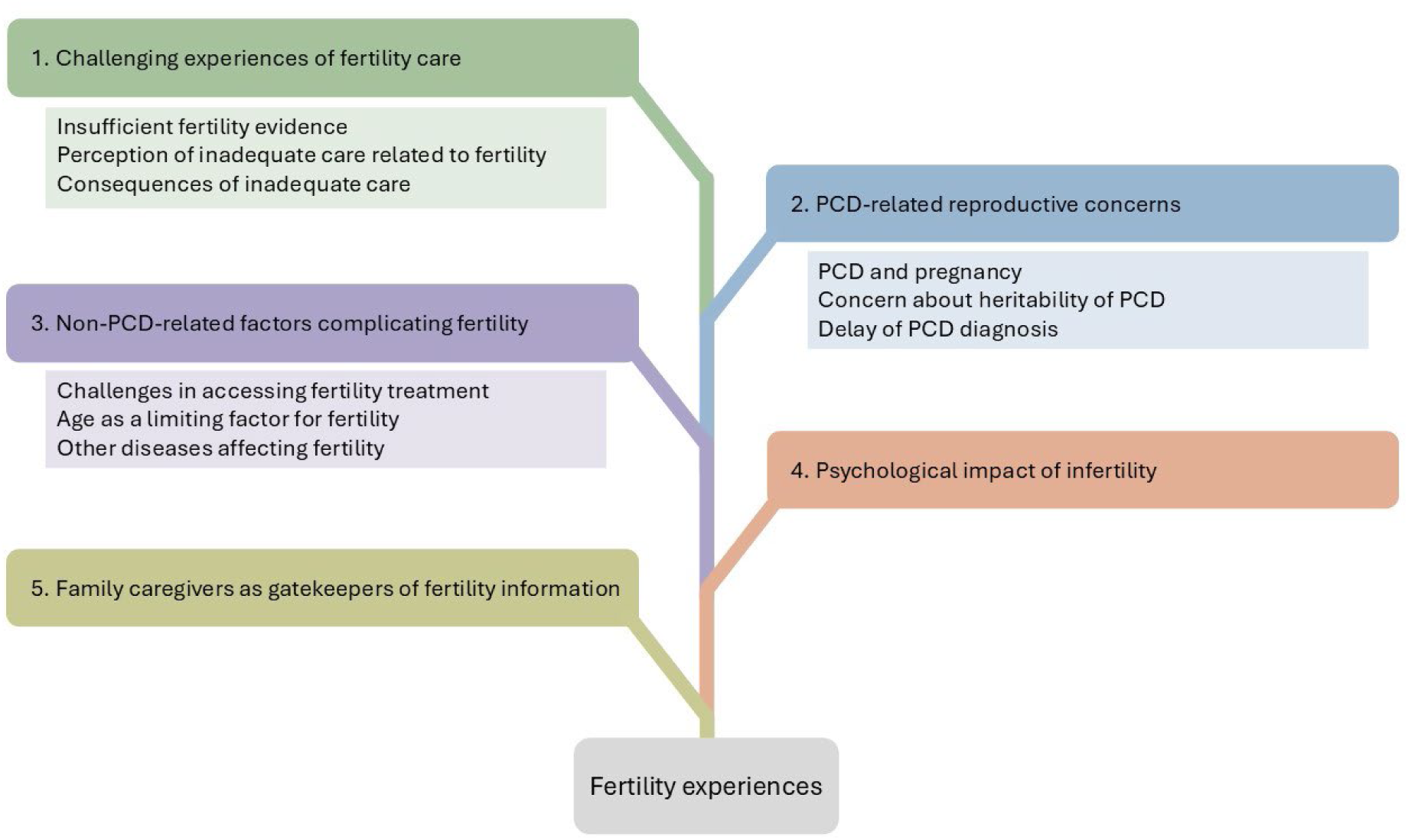
Categories related to the experiences with fertility of people with PCD and their family caregivers in the *Living with PCD* study.

## 1. Challenging experiences of fertility care

Participants reported different challenging experiences ranging from *insufficient fertility evidence* to poor care from their respective doctors leading to an overall *perception of inadequate care related to fertility*. The *consequences of inadequate care* were manifold: The perception of inadequate care led to the feeling that trying to conceive naturally was a “waste of time” rather than starting fertility treatment early. The challenging experiences motivated some participants in self-advocacy efforts to organise, research, reach out to each other and exchange experiences. Still, some participants reported having had good experiences and good care, albeit in a minority of cases.

### Insufficient fertility evidence

Several participants detected a lack of sufficient scientific evidence on fertility in PCD. In describing this deficiency, they stated it was “really hard to find the information” needed on this topic. They acknowledged that not everybody with PCD was infertile but expressed confusion and uncertainty about how PCD affects fertility and pregnancy. Since fertility problems in PCD are not easily predicted but many anticipated difficulties getting pregnant, participants also reported unexpected fertility experiences:

> “However, without looking for it, I unexpectedly became pregnant at the age of [30-39]. My pulmonologist at the time called me ‘miraculous’ and concluded that, fortunately, medicine does not explain everything.”.

> *(P181, woman, ≥ 50 years, with one child)*

Some stressed that it was difficult not knowing if people with PCD were fertile:

> “Isn’t it psychologically tricky not to know if you are fertile? Also, if you think you might not be, as a teenager, you might become more careless about protection?”.

> (P167, parent of a child, 0-5 years)

In several cases, participants hypothesized that fertility was not a priority in PCD care and research. Indeed, they felt that there is “not a huge amount of urgency for better understanding these issues in the medical community”. Adding to this, participants postulated that information was particularly lacking for women, leaving them with many open questions.

### Perception of inadequate care related to fertility

Looking at care provision, many participants were not satisfied with the care provided by their treating physician, be it a PCD physician or a fertility expert. Most reported a lack of communication with the PCD team about fertility. Many had not even been informed about possible fertility issues until late adulthood:

> “I have always been amazed that nobody has ever spoken to me about PCD and fertility issues, despite being diagnosed in my early 20s by a leading PCD hospital and being treated by them ever since. I am now beyond the age of having children, but I would hope this is something that will be improved in the future for younger generations.”.

> (P44, woman, ≥ 50 years, has never tried to become pregnant)

In some instances, participants even considered the information received as wrong: for instance, that “a PCD patient is by definition infertile” or “people with PCD are […] sterile”. Participants also pointed out that fertility experts lacked knowledge about PCD, which prevented them from effectively addressing their specific information needs.

Further, participants perceived the way fertility information was disclosed as challenging due to physicians’ lack of empathy. They reported that both PCD physicians and fertility specialists often provided this information in an offhanded manner, without sufficient explanation or context.

Participants often felt like they had “to prove” that their infertility was caused by PCD and fight for their fertility treatments. In several cases, participants stated that their physicians did not believe or listen to them. They mentioned that their fertility specialists sometimes blamed their fertility problems on other conditions. Some also commented that they were not offered fertility treatments despite their PCD diagnosis:

> “The first fertility doctor I saw refused to accept that [PCD] could be impacting on my fertility and wouldn’t accept me for in vitro fertilization (IVF) treatment […]”.

> (P31, woman, ≥ 50 years, with one child conceived via fertility treatment)

Thus, participants underlined the need for fertility education and counselling for health professionals on PCD.

### Consequences of inadequate care

A perception of “wasted time” was often mentioned as a consequence of perceived inadequate care: Long periods of trying to conceive naturally led some participants to feel that they wasted time due to bad fertility care. They were convinced that, to conceive successfully, they should have gotten information and support much earlier. This experience left many participants frustrated:

> “I was actually very frustrated in the end that the doctors didn’t give me any information at all, otherwise I would have tried to have a child much sooner.”.

> (P165, woman, ≥ 50 years, with one child conceived via fertility treatment)

As a consequential effect, some participants exhibited considerable self-advocacy motivated by the challenges they faced, and thus felt themselves responsible for seeking appropriate care. They reported managing their fertility journey without much support from health professionals and resorted to finding support and information from peers, trusting to know their body, and coming up with their own explanations regarding fertility.

> “I had the usual treatment from a professor of pneumology, an ENT specialist, a gynaecologist, and a [general practitioner]. None of these health professionals alerted me to the possibility of having difficulty conceiving a child naturally. I therefore took the personal step of calling a [fertility] centre but with a certain fear of not being legitimate, given that PCD is little known by health professionals.”

> (P190, woman, 30-39 years, pregnant at the time of the survey)

## 2. PCD-related reproductive concerns

In addition to the effects of PCD on fertility, participants shared other reproductive concerns arising from having PCD, such as *PCD and pregnancy*, particularly the impact of PCD on a potential pregnancy or on their ability to care for children, as well as *concern about the heritability of PCD*. Some participants were not even aware they had PCD when they were pregnant or faced fertility problems and this *delay of a PCD diagnosis* hindered optimal care.

### PCD and pregnancy

Some women perceived pregnancy as a health risk, either because of negative information from their PCD physicians or because of issues they had already experienced, such as ectopic pregnancies. One woman reported that her physician advised against a pregnancy due to poor lung health. Others did not even consider pregnancy because they felt they were too ill to look after children due to PCD and were already struggling to look after themselves:

> “I ultimately decided I was already too sick to care for children and to not try, which was/is heartbreaking.”.

> (P164, woman, 30-39 years, has never tried to become pregnant)

Some women who became pregnant reported that PCD affected their lung health negatively:

> “The whole pregnancy and birthing process affected my lungs and respiratory wellbeing in a negative way very considerably, […] and it would have been a much better experience and treatment outcome for me if there had been a combined treatment approach from respiratory and fertility/obstetric specialists.”.

> (P110, woman, ≥ 50 years, with two children)

Interestingly, however, some women reported better lung health during their pregnancy, providing “highest lung function results” or stating that “lung health was overall unusually good” during their pregnancy:

> “Oddly enough, while I was pregnant I was at my healthiest. My lungs and infections were the best they have ever been.”

> (P112, woman, 40-49 years, with two children)

> “I find it much easier to clear mucus when pregnant - if only my lungs could stay that way all the time!”.

> (P41, woman, 40-49 years, with two children)

### Concern about heritability of PCD

Many participants were concerned about passing PCD on to their children and wanted to better understand and be informed about the risk of the heritability of PCD. They highlighted the need for genetic counselling before getting pregnant to understand the risks of passing PCD on to their children. One person decided against having children due to the probable inheritability of PCD:

> “[…] the idea that I, as a person with a genetic disease, could pass on the corresponding genes to my offspring is a weighty point for me in my (at least current) attitude of not wanting to have children.”.

> (P94, man, 30-39 years, has never tried to father a child)

### Delay of PCD diagnosis

The cause of their fertility issues remained unclear for several participants due to delays in their PCD diagnosis. While trying to get pregnant, they had not yet been diagnosed with PCD which made optimal care more difficult. For some, it was not until years later, when PCD was diagnosed, that they finally had an explanation for their infertility. In other cases, fertility problems even raised the suspicion of PCD, and they were diagnosed shortly after they encountered fertility problems:

> “Infertility (no sperm at all in the spermiogram) was clarified twice, as the attending doctor assumed a test error. The diagnosis of PCD was also suspected a few months later due to the infertility by a general practitioner who referred me for clarification.”.

> (P1, man, ≥ 50 years, never fathered a child)

## 3. Non-PCD-related factors complicating fertility

Apart from these PCD-specific factors, participants mentioned other fertility challenges complicating their fertility journey. Among these, *challenges in accessing fertility treatment*, *age as a limiting factor for fertility* and *other diseases affecting fertility* constituted the main themes.

### Challenges in accessing fertility treatment

Availability and access to fertility treatment itself were described as an additional challenge, also in relation to the burden of the procedure. Not all participants who had problems conceiving were able to access fertility treatment due to financial or legal issues, or due to limited availability in their country at the time they were trying to conceive. One participant reported that he was only able to access fertility treatment after moving to another country because he was not entitled to it in his home country. But even those who were able to receive treatment reported ongoing difficulties, as accessibility was only the first hurdle. Many participants who underwent fertility treatment reported long and difficult journeys with many negative results until they managed to conceive a child. Fertility treatments were often described as stressful and emotionally difficult. Participants described the long duration and limited success rates of fertility treatments with many unsuccessful attempts.

> “Although I had two babies with the help of IVF, it doesn’t tell the whole fertility treatment story. In total, I went through three different rounds of fertility treatments with IVF. In all three rounds combined, I had five chromosomally normal embryos, which we attempted to transfer. Two never took, one was an early miscarriage, and two were my precious babies who are now [0-5 years] and [0-5 years] old. Prior to IVF, we also tried timed intercourse with the help of ovarian stimulation, as well as intrauterine insemination over the span of a year. And, of course, there were the two years spent trying on our own to get pregnant before we consulted with fertility doctors.”

> (P113, woman, 40-49 years, with two children conceived via fertility treatment)

Some participants decided against fertility treatments due to concerns with the technology itself. “There is a life without children, and I don’t like this whole fertilisation technology.” (P101, woman, ≥ 50 years, has never been pregnant)

### Age as a limiting factor for fertility

It is important to note here that PCD was not the only factor affecting fertility. Mainly women reported that they were aware of age as a limiting factor for fertility. Yet, some stated that they were not made aware by their treating physician that age constitutes a success factor for fertility treatments too and that an early start is important due to the long duration of treatments. Some, therefore, regretted that they had not been offered fertility treatment sooner. Participants in their late 30s or 40s who were not successful in conceiving a child described that they felt rushed because they felt time was running out and did not want to waste any more time.

> “I’m on the waiting list for IVF, which might take up to two years’ time, so I regret I didn’t start the process earlier because I’ll turn [30-39] by then.”

> (P17, woman, 30-39 years, has never been pregnant)

### Other diseases affecting fertility

Some participants–mainly women– suspected or knew that other diseases or conditions in addition to PCD, such as polycystic ovaries, irregular periods, endometriosis, or varicocele, could affect their fertility or that of their partner, which complicated their fertility journey.

> “My partner does not have PCD but does also have some fertility challenges […] and this was relevant to our journey.”

> (P21, man, 30-39 years, has never fathered a child)

## 4. Psychological impact of infertility

Infertility was described as an emotionally difficult topic and as having a huge impact on participants’ lives. One person described infertility as “the hardest part of having PCD”.

> “Fertility has been a much bigger issue for me with PCD than any lung problems, although I have a significant wet cough daily.”

> (P170, woman, 30-39 years, with two children)

Participants who managed to have children with or without the use of fertility treatments reported that they felt lucky and did not take their children for granted. They reported feeling grateful for their children and used the term “precious” when referring to the children they had managed to conceive with the help of fertility treatments. Other participants were unable to have biological children and suffered due to infertility and childlessness.

> “I am now [40-49] years old and have given up of course. It just wasn’t in my stars to become a mother.”

> (P146, woman, 40-49 years, has never been pregnant)

> “I will never have a family. Such heartache. I hope things improve for people with PCD.”

> (P158, woman, 40-49 years, has never been pregnant)

The strategies for coping with involuntary childlessness varied. Some reported that they had come to terms with the situation and showed emotional acceptance. For them, a life without children was also worth living. Others turned to adoption, became foster parents or used donor oocytes/sperm as alternative plans to parenthood.

> “[…] we were blessed with the opportunity of adopting a […] baby. She is now [30-39] years old. We were both happy with this decision and did not have the need to continue with fertility treatments.”

> (P171, woman, ≥ 50 years, has never been pregnant)

## 5. Family caregivers as gatekeepers of fertility information

Family caregivers of children with PCD not only felt responsible for their child’s current health but were also concerned about possible future health problems, of which they believed fertility was an important aspect.

Many family caregivers stated that they were aware of possible fertility problems, and most felt it was important that they were informed at diagnosis that their child might have fertility problems. However, family caregivers assessed their own need for information and that of their children differently as many did not see the need to inform their children about fertility at an early age. By doing so, they acted as gatekeepers for fertility information. Several family caregivers of children with PCD wanted to be the ones to inform their children before the physicians did and reinforced their responsibility to determine the right time to educate their child about possible fertility problems.

However, they had different perceptions of when the appropriate time for this information was. Some family caregivers had already informed their child about possible fertility problems early because they felt that this was important information for their child’s future planning:

> “As a parent of children with PCD, I think that if they are going to have issues with fertility, they should know earlier in life. It’s part of planning their futures. They should keep in mind the goals and the obstacles they will need to overcome.”.

> (P80, parent of a child, 6-12 years)

Others only wanted to inform their children when they were “adolescents/young adults”, “during puberty or later when she grows up or becomes pregnant” or “when the time comes that he wants children”.

Many were worried about how their child would perceive this potentially distressing information. One mother even indicated that she was completing the survey for her teenage son:

> “I’m a mother of a teenager with PCD, and I completed that particular survey on behalf of my son. Every survey in the past was completed by him in person. This one I did by myself because we have not talked to our child about fertility yet. His doctor says we should not worry until we have results of sperm tests and for now, he is too young for it. So we wait. But I think that [the] fertility topic is very important and every patient should be fully informed.”.

> (P64, parent of an adolescent, 13-17 years)

Although they wanted to determine the timing of the information themselves, many family caregivers expected their child’s physician to provide all the important information afterwards. Some relied on the fact that healthcare professionals should and will be available to provide fertility information when they need them to discuss this information:

> “[…] knowing that at the right and appropriate time, they can have access to and be provided with specialist counselling and help is the key. They are not alone to find out the information by themselves.”.

> (P24, parent of a child, 6-12 years)

Many described the importance of gradual, age-appropriate information, with fertility counselling only being offered to adults.

## Discussion

People with PCD reported challenging experiences related to their fertility. They faced uncertainty and disjointed care when making family planning decisions and were often left to navigate their fertility journey on their own with little support from healthcare professionals. Reproductive concerns among women about the impact of a potential pregnancy on their lung health and fear of passing on PCD unsettled them in their decision to have children. Additionally, challenges in accessing fertility treatment, the time-critical nature of fertility, and the long delays to receive a PCD diagnosis complicated their fertility journey. Infertility was described as a significant psychological burden, which was also recognised by family caregivers of young children.

This is the first study to explore the experiences of people with PCD regarding fertility. The large sample size and international scope of our study allowed us to capture insights from a diverse group of participants worldwide. The open-ended comment box in our questionnaire enabled participants to share a wide range of experiences, which resulted in a rich and varied collection of perspectives from individuals with PCD and their family caregivers. Additionally, the anonymity of the survey may have led them to share their experiences more openly.

The use of questionnaires as our data collection method limited the depth of information we could obtain, as this approach did not allow us to ask probing or follow-up questions, as would be possible in an interview. Another key limitation of this study is the lack of purposeful sampling, as participants were self-selected and completed the questionnaires based on their own interests. It is likely that individuals with fertility issues were more inclined to participate, and those with negative experiences may have been particularly motivated to share their experiences. It is also important to note that the experiences shared cover a long period of time during which significant advances in knowledge and care have been made. Participants’ reflections may, therefore, reflect historical gaps that have been partially addressed in recent years. Additionally, our study primarily reflects the experiences of people with PCD who are in contact with PCD support groups, predominantly from high-income countries in Europe and North America, where also PCD diagnosis and care are more established. Unfortunately, our study does not include individuals from minority groups where cultural identity is often closely tied to motherhood (Manouchehri et al., 2024) and we could not capture sociocultural aspects related to infertility. Infertility can carry a significant stigma, and societal norms, cultural expectations, and personal experiences strongly influence the path to parenthood.

Our study shows that people with PCD often have challenging experiences with fertility care. Many are not treated in PCD centres but by physicians with little experience with the disease, a difficulty shared with people living with other chronic diseases. People with multiple sclerosis or chronic lung diseases such as asthma also report a lack of discussions, information and education and little knowledge from healthcare professionals about fertility (Fragkoudi et al., 2023; Williams et al., 2021). Even individuals with cystic fibrosis (CF), for whom the effects on fertility are known and therefore organised care could be expected, are not satisfied with their fertility care and wish for earlier and improved fertility education (Kazmerski et al., 2017; Hailey et al., 2019). Women with CF also share concerns about the impact of their chronic condition on reproduction, as was the case with our participants. CF influences them in their pregnancy and parenting decisions as they feel they may not be well enough to support a family or conceive children (Leech et al., 2021), similar considerations have been shared by women with asthma or sarcoidosis (Williams et al., 2021). While people living with chronic diseases often become experts in managing their condition, they rely heavily on physicians and the health system for fertility issues if they need treatment. We have found that inequities in accessing fertility care further complicate the fertility journey. Additionally, for many individuals, advanced fertility treatments like IVF are not affordable because of their healthcare system (Njagi et al., 2023).

Our findings show that the combination of these diverse challenges can lead to detrimental consequences for people living with PCD who face fertility issues, particularly in terms of psychological well-being and related concerns. Research with childhood cancer survivors has shown that the psychological dimension is often overlooked in discussions about infertility. Newton et al. pointed out that survivors of childhood cancer often feel the ongoing psychological distress that results from living with uncertain fertility (Newton et al., 2021). The unpredictability of their illness worsens this burden.

Our findings also show that family caregivers are aware of potential fertility issues but often become gatekeepers of this information about their child’s fertility and future reproductive health, at times deliberating about the best timing to deliver this information. Family caregivers of children with PCD had previously expressed fertility concerns about their child’s future, worrying about the impact of fertility treatments and whether PCD could be passed on to grandchildren (Driessens et al., 2022).

However, by acting as a gatekeeper, parents themselves may limit children’s ability to receive optimal care. Shifting responsibility to adult care may carry the risk of important knowledge being lost as care for adults is less well organised than paediatric care in PCD in many countries (Lucas et al., 2014). These results point to the need for appropriate guidance for parents of children with PCD in how to best hold these discussions with their children, as well as about the appropriate timing to raise the topic.

We also found that people with PCD have a high need for information and support in relation to their fertility. This need is particularly pronounced among those who are not treated in specialised PCD centres and instead rely on physicians with limited experience in treating the disease. They wish for clearer information from healthcare professionals, more information about the impact of the disease on fertility and better integration of fertility and respiratory services. To improve their care, we need to improve the existing evidence base on fertility and create disease-specific guidelines to provide them with accurate information and counselling on their fertility journey. One participant mentioned that leaflets on PCD and fertility would be useful to share with general practitioners and other health professionals. Others mentioned that sharing studies, statistics and fertility solutions for people with PCD within the community would be very useful and appreciated. People with PCD would benefit if the psychological aspect of fertility was better integrated into their care. Information about fertility needs to be communicated honestly, sensibly, and in a timely manner, recognising concerns and offering support. By being aware of the risk of psychological effects of fertility and infertility problems, healthcare professionals can also identify those who need support and offer them professional counselling, for example, by a psychologist or social worker. Above all, accurate and timely PCD diagnosis is important to ensure that those affected receive appropriate care and fertility information.

We need enhanced support and standardised reproductive health care for people with PCD and their family caregivers to enable informed decisions on fertility, as well as to reduce the fertility-related concerns and psychological impact faced by many of them.

## Data availability

*Living with PCD* data is available upon reasonable request by contacting Claudia Kuehni.

## Ethics approval and consent to participate

Bern Cantonal Ethics Committee (Kantonale Ethikkomission Bern) approved our study (Study ID: 2020-00830). Participants provide their informed consent to participate in the study at registration. Participation is anonymous. Participants can withdraw consent to participate at any time by contacting the study team.

## Acknowledgements

We thank all participants and their families, and we thank the PCD support groups and physicians who advertised the study. We thank our collaborators who helped set up the *Living with PCD* study: Cristina Ardura, Christina Mallet, Helena Koppe, Dominique Rubi from the University of Bern and Jane S Lucas and Amanda Harris from the University Hospital Southampton. We thank Sophie Christin-Maitre (Sorbonne University), Bernard Maitre (Université Paris-Est Créteil), Nathalie Massin (American Hospital of Paris), and Lara Gonçalves Pissini (University of Bern) for their help in the design of the fertility questionnaire.

## Declaration of generative AI and AI-assisted technologies in the writing process

During the preparation of this work the author(s) used ChatGPT 4o, DeepL, and Grammarly in order to improve the readability and clarity of the manuscript. After using these tools/services, the author(s) reviewed and edited the content as needed and take(s) full responsibility for the content of the publication.

## Authors’ contributions

LD Schreck, ESL Pedersen, CE Kuehni, and M Goutaki made substantial contributions to the study concept and design. LD Schreck, ESL Pedersen, CE Kuehni, and M Goutaki were involved in the design of the fertility questionnaire. LD Schreck and S Meyer analysed data and drafted the manuscript. LD Schreck, S Meyer, ESL Pedersen, YT Lam, H Silberschmidt, S Bellu, S Zambrano, CE Kuehni, and M Goutaki critically revised and approved the manuscript.

